# Positioning Early Phase CNS Trials for Regulatory and Investor Success: Strategic Implications of the Single Phase 3 Approval Paradigm

**DOI:** 10.64898/2026.06.05.26353604

**Authors:** Peter Schmidt, Sheldon Preskorn

## Abstract

In February 2026, the FDA announced that a single pivotal phase 3 (P3) trial would become the new default standard for drug approval—a regulatory direction that had been legally enabled since the FDA Modernization Act of 1997. This announcement has strategic, scientific, and economic implications for drug developers, contract research organizations (CROs), and biotech investors. We argue that the expansion of this framework, originally reserved for various niche submissions, represents a paradigm change, dramatically increasing the value of rigorous early phase (P1 and P2) trial design, requiring sponsors to establish both statistical efficacy signals and mechanistic biological understanding before entering phase 3. Using a CNS indication cost model, we show that single P3 approval can reduce total development expenditure from approximately $447 million over 14 years to $297 million over 12 years—a savings of $150 million and providing two years of additional commercial runway for a modeled CNS drug. Case examples including lecanemab, omaveloxolone, and tofersen illustrate how biomarker-informed early phase strategies can establish the confirmatory evidence necessary for single-trial approval. We provide practical guidance for maximizing the value of P1 and P2 under this evolving framework.

## I. INTRODUCTION

### 1.1 Regulatory Background

The traditional framework for pharmaceutical drug approval in the United States has long been anchored in the requirement for two adequate and well-controlled clinical investigations demonstrating substantial evidence of effectiveness. Although the requirement for two trials was not specified in the regulations [1], it was sufficiently adopted as an FDA standard that, after the 1997 FDA Modernization Act specifically authorized approval on the basis of a single phase 3 trial [2], FDA officials indicated in 2004 that this would happen “rarely” [3]. This two-trial standard, which emerged from an era characterized by limited biological understanding of disease mechanisms and relatively crude pharmacological tools, was designed to provide statistical redundancy as a safeguard against false-positive approvals.

However, the scientific landscape of drug development has transformed profoundly over the past three decades [4]. Modern drug development now benefits from validated biomarkers, high-resolution imaging, advanced genomics, precise pharmacokinetic/pharmacodynamic (PK/PD) modeling that covers not just blood concentrations but can include variation in drug concentrations within on- and off-target tissues, and a far deeper understanding of disease pathophysiology at the molecular level. It is empirically clear that the two-trial requirement has increasingly been viewed by leading regulatory scientists as a legacy of a prior era—one that imposes substantial costs and delays without commensurate gains in evidentiary certainty when the underlying biology is well understood and characterized [5].

Despite statutory authorization, industry practice largely continued to default to the two-trial framework for novel molecules except in special circumstances such as Orphan Drug Designation, Accelerated Approval Pathway, Breakthrough Therapy Designation, Fast Track and Priority Review.

FDA guidance changed materially in February 2026, when FDA leadership announced in an article in the New England Journal of Medicine that a single pivotal trial would be the new default standard for drug approval [6]. This announcement shifted agency posture to alignment with several prior guidance documents supporting a single pivotal trial strategy [5]. If this new posture holds, it will represent a decisive inflection point for the strategy and economics of drug development and for the competitive positioning of companies capable of designing early phase studies based on robust preclinical evidence.

Moving forward, sponsors, investors, and CROs involved in early phase biopharma will need to consider the strategic implications of the new default for drug developers. What follows offers a practical framework for how early phase trials should be designed to capture the full value of this regulatory evolution.

### 1.2 The FDA Modernization Act of 1997

Section 115 of FDAMA amended Section 505(d) of the Federal Food, Drug, and Cosmetic Act (21 U.S.C. 355(d)) to provide explicitly that, where the Secretary determines—based on relevant science—that data from one adequate and well-controlled clinical investigation plus confirmatory evidence are sufficient to establish effectiveness, such data may constitute substantial evidence. This statutory language was not widely operationalized in the years following enactment, but it established the legal architecture and initiated the consideration and planning for the new approach.

Over the intervening years, the FDA issued guidance documents further elaborating on how single-trial approval could be achieved. Guidance documents including 2019 [5] and 2023 [7] publications articulated the pathway to this accelerated approval.

### 1.3 FDA Criteria for Single-Trial Approval

Under the framework established by FDA guidance and consistent with FDAMA, the agency considers the following factors in evaluating whether a single adequate and well-controlled clinical investigation plus confirmatory evidence is sufficient:

- The persuasiveness of the single trial, including internal validity, statistical rigor, and effect size magnitude;
- The robustness of the confirmatory evidence, which may include data from related populations, class evidence from pharmacologically similar agents, real-world evidence (RWE), or strong mechanistic data;
- The seriousness of the disease, particularly in settings of unmet medical need;
- The size of the relevant patient population; and
- Whether it is ethical and practicable to conduct more than one adequate and well-controlled clinical investigation.

Several of the categories of confirmatory evidence presuppose a drug that patients may encounter outside of a trial setting, for example molecules encountered in nature or drugs in trials or approved for a different indication that may provide insight into efficacy. These may result in useful trial or real-world evidence or may supplement control group data. However, in the case of a novel molecule, the primary pathway to deliver confirmatory evidence prior to FDA’s approval of the P3 strategy – the point where sponsors receive approval to plan for NDA submission after a single, successful pivotal trial – is through well planned analysis of efficacy signals identified based on a biological hypothesis developed during preclinical research and supported through data collected during phases 1 and 2.

FDA guidance states that sponsors should consult with FDA in advance of designing P3 studies under this framework. This pre-P3 engagement is not merely procedural—it is strategically essential, as the agency’s agreement on what will constitute adequate confirmatory evidence must be established before the confirmatory trial commences.

### 1.4 The February 2026 NEJM Announcement

The February 2026 announcement by FDA leadership that a single-trial requirement would be the agency’s new default standard represents not a change in law, but a shift in regulatory culture and expectation. As the Prasad and Makary commentary in the New England Journal of Medicine articulated, the prior reliance on two clinical trials was designed to provide credible causal evidence of treatment benefit in a world where biological understanding was more limited than it is today. The modern drug development toolkit—encompassing validated biomarkers, mechanistic understanding of receptor pharmacology, and sophisticated adaptive trial designs— renders the statistical redundancy of the two-trial framework less necessary when it is paired with deep biological characterization.

The announcement was widely noted by the legal and investment communities. ArentFox Schiff observed that the new default standard may offer opportunities to streamline development timelines and reduce costs, while noting that companies should expect rigorous scrutiny and will need to engage early with FDA on trial design, control groups, and confirmatory evidence strategies [8]. Gibson Dunn & Crutcher highlighted the transactional implications: milestone definitions in licensing and collaboration agreements, diligence frameworks for mergers and acquisitions, earnout structures, royalty purchase agreements, investor models, and SEC risk factor disclosures all require reconsideration in light of an approval pathway that may be materially shorter and less capital intensive than previously assumed [9].

## II. Methods

### 2.1 Development Cost Model: CNS Indication

To quantify the economic impact of the paradigm shift, we modeled clinical development costs for a representative CNS indication under both the traditional two-trial framework and the single P3 approval pathway. The model tracks annual and cumulative expenditures across the full development lifecycle, from discovery through P3 completion.

A model based on published generalized estimates for clinical development costs was created. A timeline for clinical development was created based on a review of prior analyses of phase length in the pharmaceutical development process [10, 11, 12]. Costs per phase were based on published economic analysis of these factors, including typical recruitment and duration [11,13,14,15]. CNS-specific adjustments to costs were implemented [13, 16]. For modeling enhanced trials to target the biological understanding goal in the FDA criteria in the single P3 pathway, a published approach to minimize costs in Alzheimer’s trials while incorporating imaging and fluid testing was added to the single P3 pathway for phase 1 and 2 [17].

The resulting model adds each phase together into a sequence. No modeling of variability of time between phases was included to address the possibility of fundraising delays or regulatory process for addressing results of prior phases. In our experience, this is an optimistic set of assumptions, however incorporating these interphase delays is challenging. We assumed that the optimal case for each scenario would be illustrative.

### 2.2 Financial Details

Specific components of the model for each phase follow. Detailed cost estimates are provided in the supplemental tables (Tables ST1 and ST2). All costs were adjusted for inflation from original survey dates and are presented in 2026 U.S. Dollars. The timeline for both models is also included (Table ST3).

#### 2.2.1: Discovery and target identification

This phase includes the costs of scientific personnel [11, 18], laboratory costs [14], safety and PK/PD modeling [19], patent filings [20], and overhead and facilities costs [11].

#### 2.2.2: Preclinical (IND-enabling studies)

This phase includes costs for personnel, pharmacology and toxicology studies [21], ADME/PK studies, animal acquisition and boarding, and bioanalytical lab costs [11, 14, 21]. In addition to this, there are increasing costs for CNS-specific mechanistic studies [5, 22, 23]. IND preparation and regulatory submission costs are also included at this stage and overhead charges are included [14]. Drawing from FDA guidance, this model reflects higher mechanistic and biomarker costs to inform clinical protocols in the single P3 pathway [5].

#### 2.2.3: Phase I clinical trials

Costs increase when modeling the progress into the clinical phases. Typically, CRO services are provided by the P1 unit however they are broken out in this model into project management, biometrics, and site and investigator costs [11, 14, 18, 34]. Costs for sophisticated tests, including CSF sampling, bioanalytics, and imaging were added, including expectations for costs of PET imaging for target engagement in the single P3 pathway [23, 24, 25, 26]. Regulatory expertise is also required for maintaining the IND and pharmacovigilance reporting [27].

#### 2.2.4: Phase II clinical trials

The P2 trial is multisite and, although site networks that provide a CRO function are an option, typically a P2 trial will have CRO functions and sites in separate entities. Clinical operations personnel, biometrics, and site costs scale more with size than complexity versus P1, as P1 trials bring their own complexity due to typically including an inpatient stay [11, 22, 24]. Working with clinical sites does increase complexity, and the single P3 workflow adds to site and site management costs [28]. CSF and imaging costs will be required in both the traditional two P3 and the new single P3 workflows, but these costs will be higher in the latter case [23, 26]. Regulatory engagement will continue [27].

#### 2.2.5: Phase III clinical trials

In P3, the two models diverge most dramatically. In the single P3 pathway, FDA agreement to this route indicates that the P2 data is aligned and the mechanism of action is aligned with the effect and shows some evidence of dose response [2, 5, 6]. This allows modeling the single P3 trial as the same or slightly smaller than each of the P3 trials in the traditional pathway. Confirmatory lab and imaging results drive the total closer to equity [25, 29]. Trial operations, including trial management and operations, investigator fees, laboratory, imaging, biometrics, pharmacovigilance, and regulatory strategy and filing costs also drive the cost totals as with prior phases, but adjusted for the P3 trial characteristics [11, 14].

#### 2.2.6: NDA and launch preparation

The NDA preparation is complex and requires the contributions of diverse experts [30]. It is also the phase of drug development undergoing dramatic change right now, and so this model should be revisited periodically [31]. NDA preparation and submission requires a team of diverse scientific experts and biometricians to support this effort [30,32]. This will require more intensive effort and cross-phase analysis in the single P3 workflow, particularly in phase IV planning [5].

Launch preparation includes activities to prepare for medical affairs and HEOR analysis to frame benefits for insurance coverage [33]. In the single P3 pathway, FDA has indicated that drugs approved on this pathway will be expected to collect “real-world evidence” post approval. Planning for this will be launch cost for this pathway [5]. Manufacturing scale-up costs can be highly variable and are not included in the model.

## III. Results

### 3.1 Development Cost Model: CNS Indication

Costs determined using this model are shown in Figure 1. Under the traditional approval pathway, the model projects cumulative expenditure of approximately $447 million over 14 years of development. The single P3 pathway, by contrast, projects cumulative expenditure of approximately $297 million completed in 12 years. This represents a reduction of approximately $150 million and a two-year compression of the development timeline.

**Figure 1.**
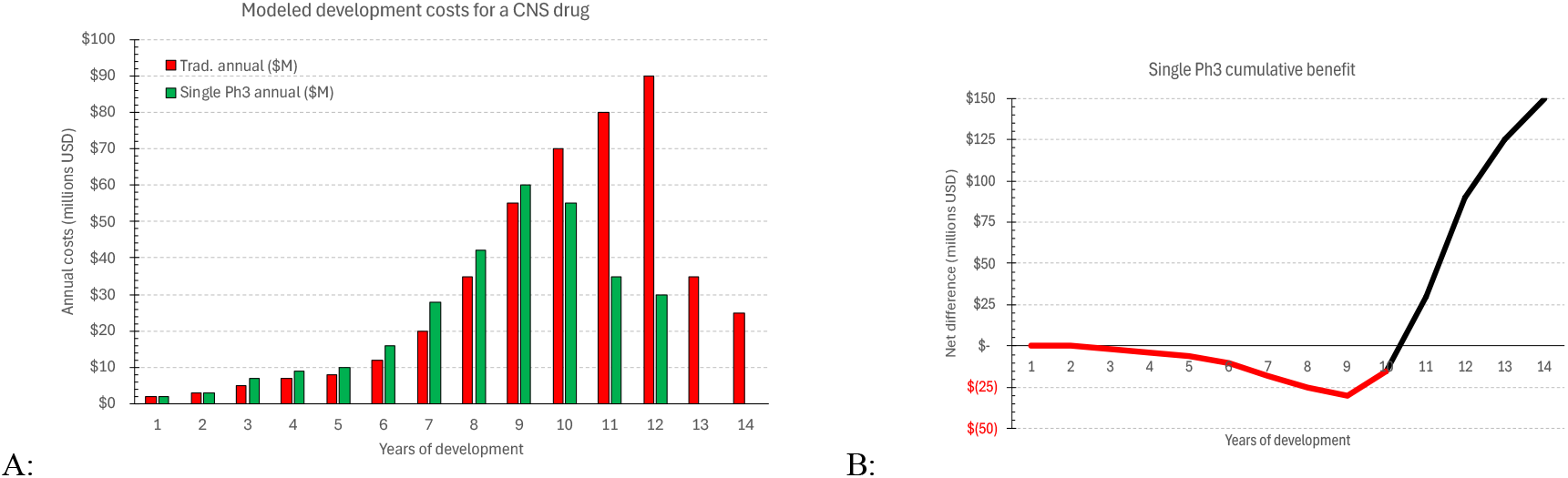
Cost models showing annual and cumulative differences. Phase endpoints: discovery, through year 2; preclinical, year 4; P1, year 6; P2, year 8; P3, year 14 for traditional and year 12 for the single P3 model.

The cost structure differences between the two pathways reflect several key features:

- Discovery: No material change in cost or timeline between pathways.
- Preclinical: The single P3 pathway requires additional investment in biomarker development and validation, modestly increasing preclinical costs. We estimate that between consulting, lab, and assay costs we expect to see an additional $4 million in costs (+33%).
- Phase 1: Enhanced PK/PD characterization, CSF sampling (for CNS indications), and biomarker integration increase P1 costs modestly relative to traditional designs. We estimate this will require $6 million in additional costs (+30%).
- Phase 2: Biomarker measurement adds cost but enables the confirmatory evidence generation that supports single P3 approval. Due to the potential requirement of nuclear imaging modalities, we estimate that this will increase the cost of P2 by $15 million (+27%).
- Phase 3: The elimination of a second pivotal trial produces the dominant cost savings. The P3 cost differential is the primary factor in savings under the single P3 pathway. We estimate that this will reduce the cost of the P3 program by $175 million (−49%).

The overall effect of the change between the traditional two P3 trial model and the modified single P3 trial with additional confirmatory evidence collected in the earlier phases is dramatic.

### 3.2 The Value of Two Additional Years of Commercial Revenue

Beyond direct cost savings, the two-year compression of the development timeline generates substantial additional value through earlier market entry. For a drug with meaningful commercial potential, two years of incremental revenue represents a compounding NPV advantage that, in many scenarios, substantially exceeds the $150 million in direct cost savings.

The paradigm shift has significant implications for drug valuations. A compound with a well-designed P1 and P2 program that demonstrates mechanistic engagement, appropriate biomarker signals, and a pre-agreed single P3 pathway has materially higher value than an equivalent compound without these features. Under the new framework, acquisition or licensing of a promising compound at the end of P2 should command a premium, reflecting: (1) the higher probability of P3 success resulting from better-informed trial design; (2) the lower expected cost of the remaining P3 program; and (3) the two-year earlier market entry. Investors and deal teams should update their valuation models.

accordingly.

## IV. Interpretation for Early Phase Trial Design

### 4.1 The “Learn and Confirm” Model

The conceptual foundation for the new paradigm is the “learn and confirm” model of clinical drug development, articulated by Preskorn and colleagues [34]. In this framework, early clinical phases (discovery, preclinical, phase 0, P1, and P2) constitute the “learn” phase—the period during which sponsors develop a deep mechanistic understanding of the drug’s pharmacology in the relevant patient population. P3 then serves as the “confirm” phase, validating at scale the hypotheses established during learning.

This is a distinct difference from the original understanding of the goals of P1, “determining the human toxicity, metabolism, absorption, elimination, and other pharmacological action…” [1]. Modern drug development often enters human trials without any human evidence for efficacy however with a solid scientific basis to anticipate it [35], a situation ideally suited for the new FDA default.

Few CNS indications have a proven biomarker for efficacy. Many novel drugs and biologics, however, target a specific mechanism under a well-supported hypothesis on the pathophysiology of the disease. A biomarker demonstrating engagement of this mechanism may be superior to a disease biomarker when combined with a significant finding of patient benefit in the P3 trial, as there have been cases of a drug modifying a disease biomarker without changing the disease [36]. Drawing on the “learn and confirm” framework, a well-executed learning phase does not simply reduce regulatory risk—it fundamentally changes the nature of P3. When the learning phase has rigorously established safety, dosing, mechanism of action, and biomarker correlates, the confirmatory trial can be designed with high precision and statistical efficiency, reducing the sample size, duration, and complexity required.

### 4.2 Three Variables Determine Clinical Response

Drug effect—and therefore clinical response—is described using Preskorn’s Organization Equation, a function of three variables whose interaction must be characterized during early phases [37]:

- Site(s) of action: characterized by the affinity and intrinsic activity of the drug at relevant receptors, transporters, enzymes, and ion channels;
- Drug concentration at site(s) of action: governed by absorption, distribution, metabolism, and elimination (ADME); and
- Underlying biology of the patient: determined by genetic factors, age, disease state, and environmental exposures (GADE).

Evaluation of these three variables during P1 and P2 from the perspective of clinical, imaging, laboratory, and pathological data (CLIP) establishes the foundation of biological safety and efficacy that FDA seeks before allowing the single pivotal trial approval pathway. Failure to characterize any of these elements adequately in early phases creates the risk of an underpowered or misdirected P3 study.

### 4.3 The Challenge of Scope in Early Phases

A central challenge in implementing the new paradigm is calibrating the appropriate scientific scope of P1 and P2. The regulatory balance is subtle: pursuing too many mechanistic questions during early phases risks generating data that the FDA may wish to see investigated further, potentially complicating the pre-P3 consultation. Pursuing too few mechanistic questions puts the “confirm” aspect to P3 at risk, with critical biological uncertainties requiring significant new findings during the P3 program.

Early phase programs need to be adapted to reflect the material shift in FDA expectations going into P3. The benefit of generating robust biological understanding during P1 and P2 is now substantially greater than it was under the two-trial framework. Sponsors and CROs should recalibrate early phase scope accordingly.

### 4.4 Enriching Phase 1 Populations

Traditional P1 trials enroll young healthy volunteers (YHV) or healthy normal volunteers (HNV) to characterize safety and PK in a population very different from those who will participate in later phases. Targeted recruitment can enhance mechanistic information generated during P1. In many novel drugs and biologics, targeted mechanisms may have measurable drug effects even in healthy populations. Lab or imaging results could inform insights to inform study design for later phases when recruiting patients. Further, measured effects could inform alternate indications for evaluation. P1 design choices to consider include:

- Age-matched cohorts: For drugs targeting conditions prevalent in older populations (e.g., neurocognitive disorders), adding cohorts in the 65-75 age range within the single ascending dose (SAD) or multiple ascending dose (MAD) portions of P1 captures population-relevant ADME and tolerability data that more closely approximates the target population.
- At-risk cohorts: Subsequent SAD or MAD cohorts can include participants with mild neurocognitive impairment, strong family history of the target illness, or relevant genetic risk factors (e.g., APOE4 homozygosity for Alzheimer’s disease), enabling early signal detection in the relevant population.
- Biomarker measurement: These enriched cohorts can incorporate measurements of relevant biomarkers to detect early pharmacodynamic signals, including changes in CSF biomarkers, neuroimaging markers, or blood-based surrogates.

A clinical example illustrates the practical value of this approach: in the development of a sublingual dexmedetomidine formulation for acute agitation, PK/PD profiling in healthy volunteers determined a much lower tolerance for the drug than was subsequently found in agitated patients with schizophrenia, with patients tolerating an order of magnitude higher plasma concentrations without adverse events. This population-specific insight was developed through an adaptive study design in a phase 1/2 trial and was essential for correct dose selection in P3 [37, 38].

Similarly, in the development of an NR2B subunit-selective NMDA antagonist for treatment-resistant depression (TRD), a dose that was well-tolerated in healthy volunteers produced dissociative events in patients with biogenic amine-nonresponsive MDD, leading to dose adjustment that preserved both efficacy and tolerability [39].

### 4.5 Biomarker Strategy in Phase 2

The critical question entering P2 is not simply whether the drug produces a clinical signal, but whether it produces a biological signal that is consistent with the proposed mechanism of action and that tracks with clinical response. Establishing this connection in P2 is the primary mechanism by which sponsors generate the confirmatory evidence required for single P3 approval.

Key principles for P2 biomarker strategy include:

- Identify a biomarker with established mechanistic relevance to the disease pathway being targeted, such that the biomarker change demonstrates mechanistic engagement.
- Demonstrate dose-dependent changes in the biomarker to provide strong evidence that the drug is the reason for biomarker change. In addition, ceiling effects (if any) on such dose dependency can be valuable in defending the P3 dose.
- Establish evidence of a correlation between biomarker change and clinical outcome to support the mechanistic hypothesis [36].
- Consider in advance the implications of an absent biomarker signal. Bayesian approaches, including adaptive trial designs, can be more effective than frequentist methods in evaluating the absence of a signal.

Several sponsors studying anti-neuroinflammatory agents have used TSPO-PET imaging to identify microglial activation. In these cases, a negative or inconclusive result is a welcome outcome, and a Bayesian adaptive design may help reduce costs for nuclear imaging.

### 4.6 Rapid P1-to-P2 Transition

The efficiency of the learning phase can be substantially enhanced by using affiliated P2 trial networks to enable rapid transition from P1 completion to P2 initiation. Traditional development often involves extended gaps between phases while sponsors regroup, analyze data, and prepare new protocols. Under the new paradigm, where time compression is a primary value driver, minimizing inter-phase gaps is strategically important. An integrated CRO with both P1 and P2 capabilities—and pre-established site networks in the relevant therapeutic area—can substantially reduce this transition time.

### 4.7 Pharmacogenomic Enrichment

As pharmacogenomics matures, it is increasingly possible to identify both patient and healthy normal subpopulations likely to produce specific biomarker signals in response to a given mechanistic intervention. The P2X7 receptor antagonist example demonstrates this principle: efficacy and safety were significantly observed in participants homozygous for a P2RX7 gain-of-function allele, suggesting that genomic enrichment of the P3 population could dramatically improve the probability of success and reduce the sample size required [40]. Ideally, such a pharmacogenomic property would be identified preclinically and would be incorporated into the P1 and P2 strategy, including targeting appropriate HNVs for the P1 safety study.

## V. Discussion: Practical Strategies for Early Phase Trial Design

### 5.1 Lecanemab: Single Pivotal Trial Success

Lecanemab (Eisai/Biogen) provides a nearly ideal approval pathway for the single pivotal trial approval process. In P2, lecanemab demonstrated dose-dependent changes in both amyloid PET and plasma phospho-tau 181 (P-Tau181), with higher doses producing greater biomarker response and low doses failing to achieve meaningful amyloid clearance. These dose-dependent biomarker signals were quantitatively correlated with Aβ42/40 ratio changes, providing mechanistic specificity. The P2 program employed a Bayesian adaptive design that allowed efficient allocation away from ineffective doses, minimizing patient exposure to subtherapeutic regimens, however they collected enough low-dose data to establish the dose dependency of the response [41].

Phase 3 (CLARITY AD) then confirmed the P2 findings at scale, demonstrating significant slowing of cognitive decline (P<0.001 at 18 months) accompanied by robust amyloid clearance. The consistency of findings from P2 to P3—with P3 essentially confirming rather than discovering the drug’s properties—represents exactly the learning-to-confirmation architecture that supports single pivotal trial approval. Lecanemab received full approval in 2024 [42].

### 5.2 Omaveloxolone: Biomarker-Informed Approval

Omaveloxolone (Reata Pharmaceuticals) for Friedreich’s ataxia demonstrates the power of dose-dependent biomarker signals in early phases. The sponsor identified an efficacy biomarker reflective of NRF2 pathway activation—the proposed mechanism of action—and designed P2 to demonstrate dose-dependent changes in this marker. The single P3 study then showed both clinical benefit and positive biomarker changes. FDA explicitly cited the dose-dependent biomarker response observed in P2 as a factor supporting expedited approval [43].

### 5.3 Tofersen: Approval Despite Primary Endpoint Failure

Tofersen (Biogen), an antisense oligonucleotide targeting SOD1 for ALS, illustrates biomarker supported approval: the drug was approved despite failing to meet its primary clinical endpoint in P3. FDA agreed to accept neurofilament light chain (NfL) as a surrogate endpoint, and approval was granted on the basis of NfL reduction. NfL is increasingly an uncontroversial disease biomarker of neuronal damage [44], and reviewers found that concordance between mechanistic endpoints at 28 weeks, the pre-specified endpoint, supported approval even though the ALSFRS-R functional scale did not reach statistical significance [45]. (They noted that the ALSFRS-R did reach significance in the open-label extension at 40 weeks [46].) This case demonstrates that when FDA has high confidence in a mechanistically validated biomarker, it can support approval even in the absence of a statistically significant clinical endpoint.

### 5.4 Aducanumab: A Cautionary Contrast

The contrast between the preceding approvals and aducanumab (Biogen) is instructive. Like lecanemab, aducanumab also targets amyloid beta, and both showed biomarker signal on PET imaging. Like tofersen, aducanumab failed to reach significance in its clinical endpoint. However, aducanumab’s development program differed from lecanemab’s in critical ways: the correlation between amyloid PET signal and clinical outcome was poor; the P3 program produced conflicting results across two trials (EMERGE and ENGAGE) that were analyzed post hoc to construct a positive narrative from one trial; and FDA approval ultimately drew on P1b data to support the efficacy claim. The result was regulatory approval under Accelerated Approval with surrogate endpoint reliance—but commercial failure, as payers and physicians were unconvinced by the evidentiary package, in part due to the complex dynamics of the approval process.

The aducanumab experience illustrates a critical principle: FDA approval is necessary but not sufficient to achieve market success. Aducanumab was approved on the basis of a post-hoc analysis of a high dose subgroup, suggesting that better dose selection in the earlier phases might have led to less controversy in the process [47]. Extracellular β-amyloid peptide is not uncontroversial as a disease biomarker in AD. Given this, and the complexities of the approval process given the therapeutic dose confusion in the pivotal trial, the commercial failure of aducanumab was a predictable risk.

## VI. Strategic Implications for Valuation

The transition to a single pivotal trial default has consequences that extend well beyond clinical development strategy. Several categories of business practice require reconsideration:

### 6.1 Milestone Definitions

Licensing and collaboration agreements commonly define milestones around P2 completion, P3 initiation, and regulatory submission. Under the new paradigm, alternative approaches to milestones may offer greater value in defining value in acquisitions or licensing. A P2 completion milestone should now be assessed not merely on whether a clinical signal was observed, but on whether the phase generated sufficient confirmatory evidence (dose-dependent biomarker response, mechanism confirmation, population-specific PK/PD) to support a single P3 design. P3 initiation milestones should specify whether FDA has agreed to the single P3 pathway.

### 6.2 Due Diligence Frameworks

M&A and licensing due diligence for late-P2 or pre-P3 assets should consider including technical assessment of early phase data packages with specific attention to: (1) biomarker strategy and mechanistic validation; (2) population-specific PK/PD characterization; (3) FDA alignment on confirmatory evidence requirements based on “persuasive” mechanistic data coming out of P2; and (4) the design features of the proposed P3 that would confirm key mechanistic signals in FDA’s framework. Assets with strong early phase biological characterization and pre-agreement from FD or a clear and compelling case for a single P3 pathway should command a strategic premium.

### 6.3 Risk Factor Disclosure

Companies operating in therapeutic areas where single P3 approval is feasible and likely should update their SEC risk factor disclosures to reflect the anticipated approval pathway, adjusted development timelines, and revised capital requirements. The standard disclosure language developed under the two-trial framework may overstate the capital required and timeline expected for programs appropriately positioned for single P3 approval.

## VII. Conclusions

The FDA’s adoption of a single pivotal trial as the new default standard for drug approval is not a disruptive regulatory break—it is the logical culmination of a trajectory that began with FDAMA in 1997 and has been reinforced by three decades of advances in biomarker science, pharmacogenomics, and disease pathophysiology. While it is not yet clearly codified in law or regulation, it is the logical conclusion of three decades of progress in biological science and regulatory progress. Over the next several years, how a declared “default” will change actual regulatory pathways and, through this, valuations of drugs in development will emerge. The recent departure from FDA of both authors of the NEJM Sounding Board article raises questions about what will come next, but the body of work leading to this suggest that cautious optimism the framework will persist is appropriate.

In some trials, particularly those where the anticipated effect size on the primary outcome is expected to be small compared to subject-to-subject variability and no biomarker for efficacy is available or has sufficient support to give confidence in regulatory acceptance, sponsors will conduct more than two clinical trials [48]. This situation is complex and technically outside of Prasad and Makary’s framework, as it anticipates clinical outcomes confirming evidence of a biological modification. For this reason, this situation is also outside of the scope of this paper.

Our model is not without limitations. We have not included confidence intervals on our phase costs as the model is meant to be illustrative rather than prognostic. The source material does not provide sufficient detail to compute variances. Some reports of clinical development costs have been largely aligned with ours [11] and others have reported higher total costs [13, 14]. We did not include launch costs in our estimates, nor did we include costs associated with failed drugs. Our hope is that our model can inform the development of business plans rather than to serve in place of a detailed clinical development strategy. Those who seek to build a budget for their clinical development will be better served to review our sources to build their own model then to attempt to adjust our model to their context.

Regardless of any speed bumps in FDA’s approach to the single pivotal trial default, Prasad and Makary’s perspective on the value of the focus on biological mechanism contributing to our understanding of clinical benefits is based on a solid history of regulatory review, as we discussed. The FDA approval process relies on the same overall clinical development framework that was codified by the FDA less than a decade after Watson and Crick discovered the structure of DNA [49]. A lot has changed in biology since then, and drug developers must submit dramatically more scientific evidence now, but in the same framework. Smarter and more strategic approaches to early-phase trials will be important in satisfying regulatory requirements for this expedited pathway, and, in the process, can incrementally de-risk the development process yielding greater confidence in acquisition and licensing contracts. The two-trial framework rewarded statistical conservatism; the single-trial framework rewards mechanistic insight. Sponsors, CROs, and investors who realign their practices to this new reality—designing early phases to generate confirmatory evidence, validating biomarker-outcome correlations, engaging FDA early on single P3 design, and structuring transactions to reflect the compressed development timeline—will capture the substantial economic and competitive advantages.

## Data Availability

All data produced in the present study are available upon reasonable request to the author.

## IX. Supplemental tables: Cost estimate tables across the clinical development timeline

**Table ST1:** Traditional, two-phase 3 trial development costs. All costs in millions USD The grand total for this modeled clinical development is $447 million. Data sources are noted in the text

| Activity | Discovery | Preclinical | Phase 1 | Phase 2 | Phase 3 | NDA & Launch prep |
| --- | --- | --- | --- | --- | --- | --- |
| Years (approximate, see timeline on Table ST3) | 1 - 2 | 2 - 4 | 4 - 6 | 6 - 9 | 9 - 13 | 13 - 14 |
| Patent filing | \$ 0.2 | \$ - | \$ - | \$ - | \$ - | \$ - |
| Discovery and scientific oversight | 2.0 | 3.5 | 0.2 | 0.3 | 0.5 | 0.1 |
| Lab and assay costs (preclinical/PK/PD/Safety) | 0.8 | 2.5 | 2.0 | 4.0 | 16.0 | - |
| Preclinical modeling | 0.3 | 6.5 | - | - | - | - |
| Regulatory strategy | - | 1.0 | 0.5 | 1.5 | 4.0 | 11.0 |
| Clinical operations management | - | - | 3.5 | 7.0 | 66.0 | - |
| Biometrics (stats, programming, DM, DSMB) | - | - | 3.0 | 9.0 | 20.0 | 4.0 |
| CMC/drug supply | - | - | 1.5 | 3.0 | 24.0 | 6.0 |
| Imaging | - | - | 0.5 | 2.5 | 10.0 | - |
| Pharmacovigilance | - | - | 0.2 | 1.0 | 6.0 | 2.0 |
| Site and investigator payments | - | - | 5.0 | 23.8 | 116.1 | - |
| Biomarker/biology validation | - | - | - | 1.5 | - | - |
| Launch preparation | - | - | - | - | - | - |
| Medical affairs and HEOR | - | - | - | - | - | 10.5 |
| Overhead | 0.7 | 2.0 | 2.0 | 4.0 | 52.0 | 3.3 |
| Total: | \$ 4.0 | \$ 15.5 | \$ 18.4 | \$ 57.6 | \$ 314.6 | \$ 36.9 |

**Table ST2:** Single phase 3 trial with biological validation development costs. All costs in millions USD The grand total for this modeled clinical development is $297 million. Data sources are noted in the text.

| Activity | Discovery | Preclinical | Phase 1 | Phase 2 | Phase 3 | NDA & Launch prep |
| --- | --- | --- | --- | --- | --- | --- |
| Years (approximate, see timeline on Table ST3) | 1 - 2 | 2 - 4 | 4 - 6 | 6 - 9 | 9 - 11.5 | 11.5 - 12.5 |
| Patent filing | \$ 0.2 | \$ - | \$ - | \$ - | \$ - | \$ - |
| Discovery and scientific oversight | 2.0 | 4.5 | 0.2 | 0.3 | 0.3 | 0.1 |
| Lab and assay costs (preclinical/PK/PD/Safety) | 0.8 | 2.5 | 2.5 | 4.5 | 8.0 | - |
| Preclinical modeling | 0.3 | 7.5 | - | - | - | - |
| Regulatory strategy | - | 1.0 | 0.5 | 2.0 | 2.5 | 12.8 |
| Clinical operations management | - | - | 4.0 | 9.0 | 29.0 | - |
| Biometrics (stats, programming, DM, DSMB) | - | - | 3.0 | 11.0 | 10.0 | 4.0 |
| CMC/drug supply | - | - | 1.5 | 3.0 | 10.0 | 6.5 |
| Imaging | - | - | 3.5 | 5.0 | 5.0 | - |
| Pharmacovigilance | - | - | 0.2 | 1.5 | 3.0 | 2.0 |
| Site and investigator payments | - | - | 5.4 | 25.2 | 49.4 | - |
| Biomarker/biology validation | - | 2.5 | 1.5 | 4.0 | - | - |
| Launch preparation | - | - | - | - | - | - |
| Medical affairs and HEOR | - | - | - | - | - | 11.5 |
| Overhead | 0.7 | 2.5 | 2.5 | 5.0 | 20.0 | 3.2 |
| Total: | \$ 4.0 | \$ 20.5 | \$ 24.8 | \$ 70.5 | \$ 137.2 | \$ 40.1 |

**Table ST3:**
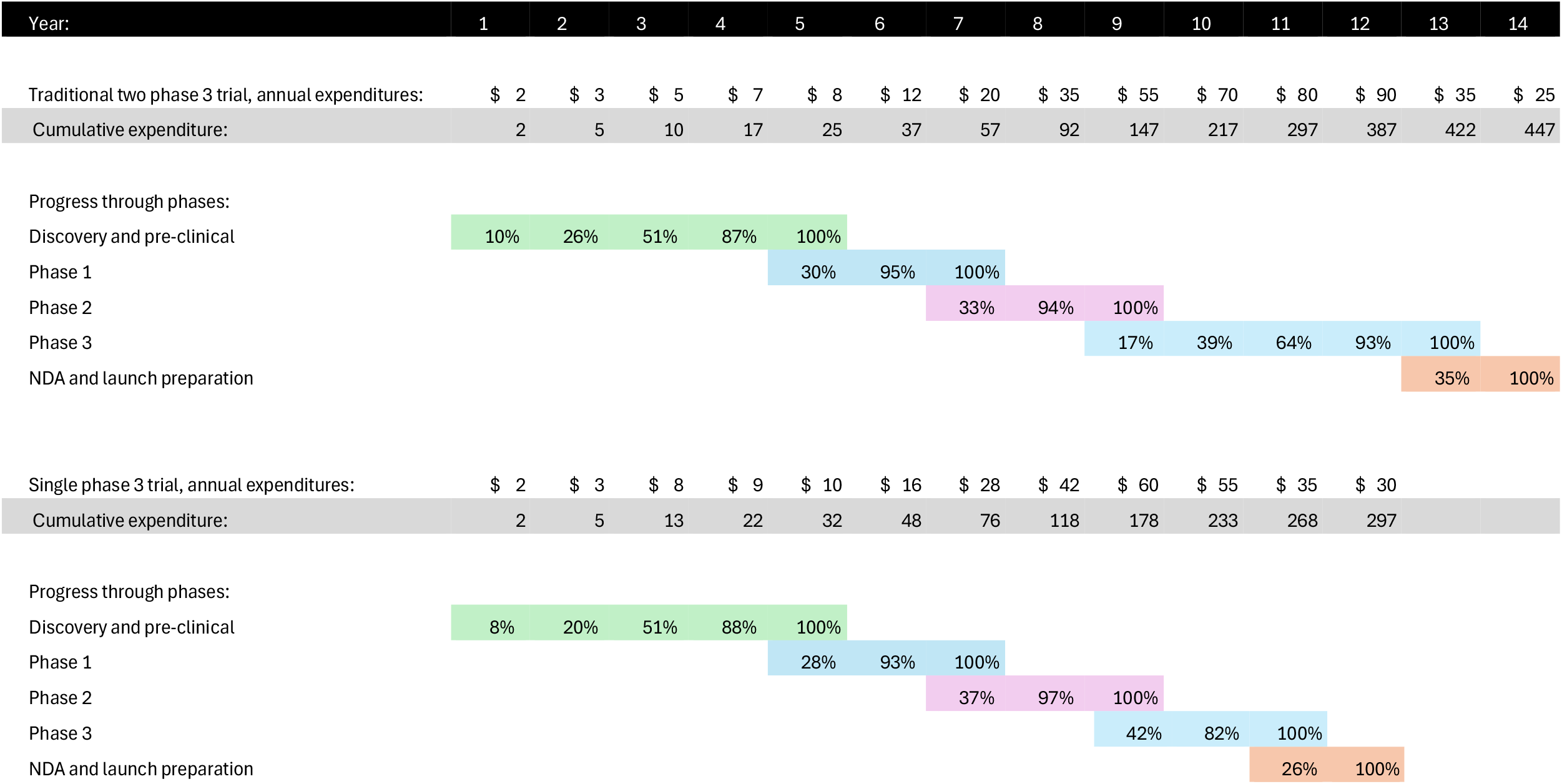
Timeline for both models. All costs in millions USD This table shows the progress through phases, with millions of dollars spent and percentage of phase complete at the end of each year in the labeled column. Note that some tail activities continue in a phase after the prior phase has started. For example, the model includes year 5 preclinical expenditures to show chronic toxicology studies ongoing to support P2 that are not required before FIH P1 studies. It is not uncommon to have trailing expenses for clinical phases, as clinical activities are closed out, secondary and exploratory outcomes are analyzed, and vendor contracts are reconciled.

## Notes

### Competing Interest Statement

The authors provide strategic guidance to companies and government sponsors developing drugs for regulatory approval.

### Funding Statement

This study did not receive any funding.

